# A protocol for a scoping review on the role of whole-body and dedicated body-part magnetic resonance imaging for assessment of adult and juvenile idiopathic inflammatory myopathies

**DOI:** 10.1101/2024.03.26.24304925

**Authors:** Mickael Essouma, Daniel Brito de Araujo, Jessica Day, Latika Gupta, Adina Kay Knight, Ann Reed, Elie Naddaf, Adriana Maluf Elias Sallum, Edoardo Marrani, Edoardo Conticini, Simone Appenzeller, Adina Kay Knight, Mazen Dimachkie, Tamima Mohamad Abou, Daren Gibson, Eva Kirkhus, Anneke J van der Koi, James B Lilleker, Matteo Lucchini, Pedro Machado, Mary Anne Riopel, Helga Sanner, Adam Schiffenbauer, Julio Brandão Guimarães, Claudia Saad-Magalhaes, Susan O’Hanlon, Clarissa Harumi Omori, Susan Phaneuf, Helga Sanner, Siamak Moghadam-Kia, Mirkamal Tolend, Iazsmin Bauer Ventura, Lisa G Rider, Lisa Christopher-Stine, Julie J Paik, Brian Feldman, Samuel Katsuyuki Shinjo, Andrea Schwarz Doria

**Affiliations:** Network of Immunity in Infection, Malignancy and Autoimmunity, Universal Scientific Education and Research Network, Yaounde, Cameroon; Internal Medicine Department, Universidade Federal de Pelotas, Pelotas, RS, Brazil; The University of Melbourne, Melbourne, Victoria, Australia; The Walter and Eliza Hall Institute of Medical Research, Melbourne, Victoria, Australia; Royal Melbourne Hospital, Melbourne, Victoria, Australia; Division of Musculoskeletal and Dermatological Sciences, Centre for Musculoskeletal Research, School of Biological Sciences, University of Manchester, Manchester, UK; Department of Rheumatology, Royal Wolverhampton Hospitals NHS Trust, Wolverhampton, UK; Department of Rheumatology, City Hospital, Sandwell and West Birmingham Hospitals NHS Trust, Birmingham, UK; Department of Clinical Immunology, Mount Sinai School of Medicine, New York, New York, USA; Department of Pediatrics, Duke University, Durham, NC, USA; Mayo Clinic, Rochester, Minnesota, USA; Pediatric Rheumatology Unit, Children’s Institute, Hospital das Clínicas, Faculdade de Medicina, Universidade de São Paulo, São Paulo, SP, Brazil; Rheumatology Unit, ERN-ReCONNET center, Meyer Children’s Hospital IRCCS, Florence, Italy; Rheumatology Unit, Department of Medicine, Surgery and Neurosciences, University of Siena, Italy; Rheumatology Unit, Policlinica, Universidade Estadual de Campinas. Campinas, SP, Brazil; The Division of Allergy and Immunology/Department of Pediatrics, Mount Sinai School of Medicine, New York, USA; Department of Neurology University of Kansas, Kansas City, KS, USA; Medical Imaging, Fiona Stanley Hospital, Murdoch, Australia; Department of Radiology, Oslo University Hospital, Oslo, Norway; Department of Neurology, Amsterdam UMC, University of Amsterdam, Amsterdam Neuroscience, Amsterdam, Netherlands; Manchester Centre for Clinical Neurosciences, Salford Royal NHS Foundation Trust, Salford, UK; UOC Neurologia, Fondazione Policlinico Universitario Agostino Gemelli IRCCS, Rome, Italy; Dipartimento di Neuroscienze, Sezione di Neurologia, Catholic University of Sacred Heart, Rome, Italy; Department of Neuromuscular Diseases and Centre for Rheumatology, University College London, UK; Department of Rheumatology and Queen Square Centre for Neuromuscular Diseases, University College London Hospitals NHS Foundation Trust, London, UK; Associate Professor, Moravian College, Bethlehem, PA, USA; Department of Rheumatology, Oslo University Hospital, Oslo, Norway; Department of Health Sciences, Oslo New University College, Oslo, Norway; Environmental Autoimmunity Group, National Institute of Environmental Health Sciences, USA; Universidade Federal de São Paulo, São Paulo, SP, Brazil; Grupo Fleury Medicina e Saúde, São Paulo, Brazil; Pediatric Rheumatology Unit, Department of Pathology, Botucatu Medical School, São Paulo State University, Botucatu, São Paulo, SP, Brazil; Patient representative; Division of Rheumatology and Clinical Immunology, Department of Medicine, University of Pittsburgh School of Medicine, Pittsburgh, PA, USA; Department of Diagnostic Imaging, Research Institute, The Hospital for Sick Children, University of Toronto, Toronto, ON, Canada; Department of Medicine, Section of Rheumatology, University of Chicago, IL, USA; Johns Hopkins Division of Rheumatology, Johns Hopkins University School of Medicine, Baltimore, MD, USA; Division of Rheumatology, The Hospital for Sick Children, Toronto, Ontario, Canada; Division of Rheumatology, Faculdade de Medicina FMUSP, Universidade de São Paulo, São Paulo, SP, Brazil

**Author notes:** **Address for correspondence:** Dr Andrea Schwarz Doria. Department of Diagnostic Imaging, Research Institute, The Hospital for Sick Children, Department of Medical Imaging, University of Toronto. 555 University Avenue, Toronto ON M5G1X8, Canada. Mickael Essouma Daniel Brito de Araujo share first authorship. Andrea Schwarz Doria and Samuel Katsuyuki Shinjo share senior authorship.

**Keywords:** Assessment, damage, idiopathic inflammatory myopathies, magnetic resonance imaging, myositis

## Abstract

**Background:** Currently, there is lack of standardization of magnetic resonance imaging (MRI) scoring systems and protocols for assessment of idiopathic inflammatory myopathies (IIMs) in children and adults among treatment centres across the globe. Therefore, we will perform scoping reviews of the literature to inform available semi-quantitative and quantitative MRI scoring systems and protocols for the assessment and monitoring of skeletal muscle involvement in patients with IIMs with the final goal of providing evidence-based information for the future development of a universal standardized MRI scoring system in specific research and clinical settings in this population.

**Methods:** Electronic databases (PubMed, EMBASE, and Cochrane) will be searched to select relevant articles published from January 2000 to October 2023. Data will be synthesized narratively.

**Discussion:** This scoping review will extensively map evidence on the indications, utility for diagnosis and assessment of disease activity and damage using skeletal muscle MRI in IIMs. The results will allow the development of consensus recommendations for clinical practice and enable the standardization of research methods for MRI assessment of skeletal muscle changes in patients with IIMs.

## INTRODUCTION

Idiopathic inflammatory myopathies (IIMs) comprise a heterogeneous group of systemic autoimmune diseases that predominantly affect skeletal muscles and can affect individuals of any age from childhood to adulthood [1,2]. Moreover, IIMs are classified based on clinical phenotype, antibody status, and post-biopsy pathological results [1-3].

Magnetic resonance imaging (MRI) has been used to evaluate IIM for decades [4]. However, there is a need for standardization of MRI techniques and protocols for body regions to be scanned (whole-body (WB) or dedicated body-part and result reports when assessing IIM-related skeletal muscle involvement. This is especially important because a vast range of skeletal myopathies share MRI features with IIMs, including edema, fat infiltration, and muscle atrophy. Evidence-based guidelines for the imaging diagnoses of IIMs are sparse.

There is a paucity of information in the literature regarding the effect of body’s growth on the imaging characteristics and signal patterns of muscles. While some muscular diseases have similar MRI appearances, others present distinct signal intensity patterns of muscle edema and fatty infiltration [5]. All these factors corroborate the fact that the current protocols for MRI data acquisition and interpretation of myopathies in children and adults differ across healthcare centers worldwide. Without standardized MRI protocols and scoring systems, it becomes challenging to compare the results of clinical trials of patients with IIMs across the globe (research setting) and for patients who are followed up in different geographically-based treatment centers to have appropriate management (clinical setting).

In addition to the standardization of MRI methodology for the diagnosis of IIM, an accurate early diagnosis is essential for clinicians to start appropriate treatment, which can prevent long-term damage.

To address the aforementioned issues, an International Myositis Assessment and Clinics Studies Group (IMACS) working group focused on MRI for the assessment of IIMs was assembled in early 2022, bringing together leading experts in pediatric and adult rheumatology and radiology, knowledge translation, stakeholders, and patients with the following goals:

a. Facilitate the future development of consensus recommendations on standardized MRI protocols and scoring systems that are most suitable for assessing skeletal muscle inflammation and damage of different muscular groups across the body in patients with IIMs of all age groups.
b. Develop evidence-based guidelines and recommendations for MRI assessment in pediatric and adult IIM, supporting future research to test the usability, reliability, and validity of the proposed standardized MRI scale.
c. To utilize artificial intelligence and machine learning tools to facilitate and augment the role of imagers and clinicians in the application of MRI scoring systems in clinical and research settings.

As a starting point for our working-group goal, we focused on the evidence-based accrual of information on MRI assessment of pediatric and adult IIMs and reviewed the recent literature on the available WB and dedicated-part MRI scoring systems and protocols for the diagnosis, assessment, and monitoring of skeletal muscle involvement in children and adults with IIMs.

The overarching questions of our scoping reviews concerning the settings (clinical practice and research) for which a future standardized MRI scoring system could be applied will be as follows, separately for pediatric and adult populations:

a. What are the available WB and dedicated-MRI techniques and protocols for assessing IIMs in children and adults in clinical and research settings reported in the literature?
b. What are the available WB and dedicated-MRI scoring systems for the interpretation and quantification of pathology in IIMs of children and adults in clinical and research settings reported in the literature?
c. Have the available MRI scoring systems undergone methodological evaluation concerning their clinimetric properties (reliability, validity, and responsiveness) [6,7]?

## METHODS

### Design

The two proposed scoping reviews are WBMRI and dedicated-MRI in a) juvenile-onset IIMs and b) adult-onset IIMs, which will follow the PRISMA extension for scoping reviews (PRISMA-ScR) guidelines [8].

### Definitions

IIMs are heterogeneous, systemic autoimmune diseases characterized by weakness, chronic inflammation of skeletal muscles, typical skin rashes (Gottron’s papules or heliotrope rash), elevated serum levels of muscle enzymes, or increased electrical activity in the muscle detected by electromyography, according to the classification criteria established by Bohan and Peter based on these features [9]. If onset occurs before the age of 18 years, it is referred to as juvenile IIM. Alternatively, if it occurs in patients aged 18 years or older, it is called adult IIM [10].

### Inclusion criteria of the scoping reviews

a. Types of studies: published peer-reviewed full-texts of observational (cases series, cross-sectional, case-control, and cohort) studies reporting MRI assessments in a minimum of five subjects with IIMs. MRI examinations will be classified according to the region of interest, as follows:
  a1) WBMRI: defined as an imaging study with a field-of-view extending from the level of the head/neck to the toes. It aims to cover the bilateral proximal muscles (shoulder and/or hip girdles), paraspinal muscles, and/or bilateral distal muscles of the upper and lower extremities and typically involves a small field of view.
  a2) Dedicated-MRI: defined as an imaging study of one body region (e.g., shoulder or hip). It includes MRI assessment of the proximal musculature of either the upper or lower extremities.
b. Participants: Human subjects with IIMs of any age, sex, race/ethnicity. The pediatric population will be defined as individuals aged < 18 years at the time of disease onset. This cutoff for age was used in other meta-analyses focused on the pediatric population [11].
c. Outcome of interest: MRI features and scoring systems used to interpret results.
d. The existence of figures that exemplify the scoring system being investigated in the primary manuscript.
e. English and non-English language (Portuguese, Spanish, French, German, Italian) in primary studies according to language knowledge of members of our Task Force. The papers will be reviewed in the native language in which they were published by 2 assigned reviewers of the Task Force according to their language expertise. These reviewers will assess the full papers for confirmation of fulfilment of inclusion criteria of our scoping review and will retrieve pertinent information as per the data charting process of our review. They will obtain consensus for data entry onto the study database.

The reviewers will assess the full manuscript to confirm if the assigned papers meet the inclusion criteria of our scoping review and will retrieve pertinent information as per the data charting process of our review.

### Exclusion criteria

The following types of articles will be excluded: abstracts of conferences, reviews, decision-making models, economic evaluations, articles evaluating etiopathogenic concepts of myopathies beyond inflammation (e.g., neuromyopathic etiopathogenesis), studies in which raw data or clarification of parts of the text remain unavailable after requesting this information from the authors, duplicated articles, salami-sliced articles [12], and other forms of plagiarized reports.

### Information sources

We will search electronic databases (PubMed, EMBASE, and COCHRANE) to identify relevant studies published from January 1, 2000, to October 15, 2023.

The rationale for the selection of this timeframe relies on the fact that upon review of primary articles on MRI assessment of IIMs before 2000, we noted that the quality of MRI (for dedicated-MRI) described in the articles was not comparable with the imaging quality of the studies published afterward. Besides that, due to the difficulty in comparing information from studies with different imaging techniques and as we intend to develop a standardized MRI scoring system, which can also benefit from the utilization of artificial intelligence and machine learning tools, we need to support our ground-based information on studies whose MRI data were acquired in MRI scanners of similar generations. Thus, we limited the timeframe of the literature search to our scoping reviews.

Detailed search strategies will be provided. Electronic searches will be supplemented by manual searches, consisting of scrutiny of references of relevant original and review articles.

### Selection of sources of evidence

Scanning of the titles and abstracts retrieved from the electronic searches will be performed independently by two investigators. Discrepancies in opinions will be resolved by consensus with other members of the group. The preliminary results of the study selection will be discussed among the Task Force subgroups through online meetings. The full texts of the selected articles will be assessed independently by pairs of investigators of the subgroups of the Task Force according to pre-established criteria (data charting process). Discrepancies among investigators regarding the evaluation of the full text will be resolved through discussion.

### Data charting process

Data will be extracted independently by pairs of investigators using a standardized data extraction sheet. The extracted data will be independently crosschecked by the convener of the Task Force subgroup and two experienced reviewers who will obtain a consensus for discrepant cases. The following data will be extracted from the included articles: last name of the first author, date of publication, study design (retrospective/prospective/ambispective), type of sampling, demographic characteristics of participants, IIM features, indications for MRI, MRI techniques used (including T1/T2/STIR sequences, WBMRI or dedicatedd-MRI, scales used to assess radiographic muscle changes, clinimetric properties of scoring systems, and methods/statistics used for testing (e.g., reference standards used for diagnostic accuracy studies), correlation between clinical-laboratory constructs (e.g., disease activity and damage measures), and MRI findings. MRI findings will include but are not limited to subcutaneous adipose tissue edema, fascial edema, distribution of muscle edema, fat infiltration, and muscle atrophy [13,14].

### Critical appraisal of sources of evidence

We will not appraise the quality of the included studies, as we will conduct a scoping review rather than a systematic review, and study quality will be an exclusion criterion for our proposed review.

### Data synthesis

We will list the search strategies for each database used in this review. The data will be qualitatively synthesized. The review will be divided into the following themes, as previously mentioned: WBMRI and dedicated-MRI for (a) juvenile-onset IIMs and (b) adult-onset IIMs.

## DISCUSSION

Our scoping review will comprehensively explore the available literature on the use of MRI for the assessment of individuals with IIMs.

The ultimate goal of our review is to develop a pediatric and an adult WB and dedicated-MRI scoring systems for the diagnosis, evaluation, and monitoring of IIM-related skeletal muscle injuries in both research and clinical practice. We hope that this review will provide detailed insights into the body of evidence for different MRI protocols and scoring methods available for IIM assessment. We further expect that the quality of evidence available will be suitable for guiding our working this IMACS Task Force towards the conduct of a survey to be applied in the global IMACS members concerning the current utilization of WBMRI and dedicated-MRI for the assessment of IIMs in research and clinical practice settings and learn about the percentage of those members who supported future guidelines on recommended MRI protocols for different purposes and settings. Moreover, our working group aims to update new knowledge on MRI assessment in IIMs to the global scientific community. The findings of this scoping review will be disseminated by members of the Task Force and IMACS through social media and websites and presented at international conferences.

We foresee some limitations of this scoping review such as the heterogeneity of the methods for reporting data across primary studies, the potential availability of papers with small numbers of participants, heterogeneous patient populations, incomplete description of MRI techniques and MRI findings, and the likely lack of representativeness of research from certain parts of the globe owing to a lack of MRI studies from some regions.

## CONCLUSION

The goal of this scoping review will be to collect data on scoring systems and protocols using skeletal muscle MRI in patients with IIMs for the future standardization of scoring methods for MRI assessment of skeletal musculature changes in children and adults with IIMs.

## Supporting information

Supplement_Material_Search_Terms

## Data Availability

All data produced in the present work are contained in the manuscript and supplementary material

## DECLARATIONS

### Authors’ contributions

Conceptualization: A.S.D., S.K.S., D.B.A., J.D., A.M.E.S., C.M., M.E.

First draft of the manuscript: M.E.

Revision: D.B.A., S.K.S., J.D., L.G., A.K.K., A.R., E.N., S.A., E.M., A.M.E.S., M.D., J.B.G., M.E. and A.S.D.

All authors participated in meetings for discussion of the scope of the review and approved the final version of the manuscript.

### Funding

This work did not receive any funding.

## Acknowledgements

The support and thoughtful comments from IMACS Scientific Committee.

## Availability of data and materials

All data pertaining to this manuscript are reported within the full text and the supplementary material.

## Declarations

JD is the recipient of the Sylvia and Charles Viertel Charitable Foundation Clinical Investigator Award, the John T Reid Charitable Trust Centenary Fellowship, the RACP Australian Rheumatology Association and D.E.V Starr Research Establishment Fellowship and the Royal Melbourne Hospital Victor Hurley Medical research Grant in Aid.

Other authors declare that they have any competing interests.

## Ethics approval and consent to participate

Not applicable.

## Notes

### Competing Interest Statement

The authors have declared no competing interest.

### Funding Statement

This study did not receive any funding

## REFERENCES

1. Lundberg IE, Tjarnlund A, Bottai M, Werth VP, Pilkington C, de Visser M, et al. 2017 European League Against Rheumatism/American College of Rheumatology classification criteria for adult and juvenile idiopathic inflammatory myopathies and their major subgroups. Arthritis Rheumatol. 2017; 69(12): 2271–82.

2. Lundberg IE, Fujito M, Vencovsky J, Aggarwal R, Holmqvist M, Christopher-Stine L, et al. Idiopathic inflammatory myopathies. Nat Rev Dis Primers. 2021; 7(1): 86.

3. Cavagna L, Trallero-Araguas E, Meloni F, Cavazzana I, Rojas-Serrano J, Feist E, et al. Influence of antisynthetase antibodies specificities on antisynthetase syndrome clinical spectrum time course. J Clin Med. 2019; 8(11): 2013.

4. Zubair A.S., Salam S, Dimachkie MM, Machado PM, Bhaskar R. Imaging biomarkers in the idiopathic inflammatory myopathies. Front Neurol. 2023; 14: 1146015.

5. Theodorou DJ, Theodorou SJ, Kakitsubata Y. Skeletal muscle disease: patterns of MRI appearances. Br J Radiol. 2012; 85(1020): e1298–308.

6. Feinstein AR, Wells CK, Joyce CM, Josephy BR. The evaluation of sensibility and the role of patient collaboration in clinimetric indexes. Trans Assoc Am Physicians. 1985; 98: 146–9.

7. Frytak J. Measurement. J Rehabil Outcomes Meas. 2000; 4: 15–31.

8. Tricco AC, Lillie E, Zarin W, O‘Brien KK, Colquhoun H, Levac D, et al. PRISMA Extension for scoping reviews (PRISMA-ScR): Checklist and explanation. Ann Intern Med. 2018; 169(7): 467–3.

9. Bohan A, Peter JB. Polymyositis and dermatomyositis. Part 1. N Engl J Med. 1975; 292: 344–7.

10. Lundberg IE, Tjärnlund A, Bottai M, Werth VP, Pilkington C, Visser M, et al; International Myositis Classification Criteria Project consortium, The Euromyositis register and The Juvenile Dermatomyositis Cohort Biomarker Study and Repository (JDRG) (UK and Ireland). 2017 European League Against Rheumatism/American College of Rheumatology classification criteria for adult and juvenile idiopathic inflammatory myopathies and their major subgroups. Ann Rheum Dis. 2017; 76(12): 1955–64.

11. Chan MW, Leckie A, Xavier F, Uleryk E, Tadros S, Blanchette V, et al. A systematic review of MR imaging as a tool for evaluating haemophilic arthropathy in children. Haemophilia. 2013; 19(6): e324–34.

12. Tolsgaard MG, Ellaway R, Woods N, Norman G. Salami-slicing and plagiarism: how should we respond? Adv Health Sci Educ Theory Pract. 2019; 24(1): 3–14.

13. Goyal NA, Mozaffar T, Dimackie MM. Imaging beyond muscle resonance imaging in inclusion body myositis. Clin Exp Rheumatol. 2023; 41(2): 386–392.

14. Day J, Patel S, Limaye V. The role of magnetic resonance imaging techniques in evaluation and management of the idiopathic inflammatory myopathies. Semin Arthritis Rheum. 2017; 46(5): 642–9.

