## Supplement_Material_Search_Terms for "A protocol for a scoping review on the role of whole-body and dedicated body-part magnetic resonance imaging for assessment of adult and juvenile idiopathic inflammatory myopathies"

**Supplementary Table 1**. Search terms from Taskforce - Group 1.

| PubMed | |
|---|---|
| **Terms** | **Strategy** |
| Inflammatory myopathies AND Whole Body Magnetic Resonance Imaging | ("myositis"[MeSH Terms] OR "myositis"[All Fields] OR ("inflammatory"[All Fields] AND "myopathies"[All Fields]) OR "inflammatory myopathies"[All Fields]) AND (("whole"[All Fields] OR "wholeness"[All Fields] OR "wholes"[All Fields]) AND ("human body"[MeSH Terms] OR ("human"[All Fields] AND "body"[All Fields]) OR "human body"[All Fields] OR "body"[All Fields]) AND ("magnetic resonance imaging"[MeSH Terms] OR ("magnetic"[All Fields] AND "resonance"[All Fields] AND "imaging"[All Fields]) OR "magnetic resonance imaging"[All Fields])) |
| Juvenile dermatomyositis AND Whole Body MRI | ("dermatomyositis"[MeSH Terms] OR "dermatomyositis"[All Fields] OR ("juvenile"[All Fields] AND "dermatomyositis"[All Fields]) OR "juvenile dermatomyositis"[All Fields]) AND (("whole"[All Fields] OR "wholeness"[All Fields] OR "wholes"[All Fields]) AND ("human body"[MeSH Terms] OR ("human"[All Fields] AND "body"[All Fields]) OR "human body"[All Fields] OR "body"[All Fields]) AND ("magnetic resonance imaging"[MeSH Terms] OR ("magnetic"[All Fields] AND "resonance"[All Fields] AND "imaging"[All Fields]) OR "magnetic resonance imaging"[All Fields] OR "mri"[All Fields])) |
| Adult dermatomyositis AND localized MRI | ("dermatomyositis"[MeSH Terms] OR "dermatomyositis"[All Fields] OR ("adult"[All Fields] AND "dermatomyositis"[All Fields]) OR "adult dermatomyositis"[All Fields]) AND (("focal"[All Fields] OR "focalities"[All Fields] OR "focality"[All Fields] OR "focalization"[All Fields] OR "focalized"[All Fields] OR "focally"[All Fields] OR "focals"[All Fields] OR "local"[All Fields] OR "localisation"[All Fields] OR "localisations"[All Fields] OR "localise"[All Fields] OR "localised"[All Fields] OR "localises"[All Fields] OR "localising"[All Fields] OR "localization"[All Fields] OR "localizations"[All Fields] OR "localize"[All Fields] OR "localized"[All Fields] OR "localizer"[All Fields] OR "localizers"[All Fields] OR "localizes"[All Fields] OR "localizing"[All Fields] OR "locally"[All Fields] OR "locals"[All Fields]) AND ("magnetic resonance imaging"[MeSH Terms] OR ("magnetic"[All Fields] AND "resonance"[All Fields] AND "imaging"[All Fields]) OR "magnetic resonance imaging"[All Fields] OR "mri"[All Fields])) |
| Adult dermatomyositis AND Whole-body magnetic resonance | ("dermatomyositis"[MeSH Terms] OR "dermatomyositis"[All Fields] OR ("adult"[All Fields] AND "dermatomyositis"[All Fields]) OR "adult dermatomyositis"[All Fields]) AND ("Whole-body"[All Fields] AND ("magnetic resonance spectroscopy"[MeSH Terms] OR ("magnetic"[All Fields] AND "resonance"[All Fields] AND "spectroscopy"[All Fields]) OR "magnetic resonance spectroscopy"[All Fields] OR ("magnetic"[All Fields] AND "resonance"[All Fields]) OR "magnetic resonance"[All Fields])) |
| Inflammatory myopathies AND dedicated MRI | ("myositis"[MeSH Terms] OR "myositis"[All Fields] OR ("inflammatory"[All Fields] AND "myopathies"[All Fields]) OR "inflammatory myopathies"[All Fields]) AND (("dedicate"[All Fields] OR "dedicated"[All Fields] OR "dedicating"[All Fields] OR "dedication"[All Fields]) AND ("magnetic resonance imaging"[MeSH Terms] OR ("magnetic"[All Fields] AND "resonance"[All Fields] AND "imaging"[All Fields]) OR "magnetic resonance imaging"[All Fields] OR "mri"[All Fields])) |
| Inflammatory myopathies AND Whole Body Magnetic Resonance Imaging | ("myositis"[MeSH Terms] OR "myositis"[All Fields] OR ("inflammatory"[All Fields] AND "myopathies"[All Fields]) OR "inflammatory myopathies"[All Fields]) AND (("whole"[All Fields] OR "wholeness"[All Fields] OR "wholes"[All Fields]) AND ("human body"[MeSH Terms] OR ("human"[All Fields] AND "body"[All Fields]) OR "human body"[All Fields] OR "body"[All Fields]) AND ("magnetic resonance imaging"[MeSH Terms] OR ("magnetic"[All Fields] AND "resonance"[All Fields] AND "imaging"[All Fields]) OR "magnetic resonance imaging"[All Fields])) |
| Autoimmune myopathies AND whole body MRI | ("autoimmune"[All Fields] OR "autoimmunity"[MeSH Terms] OR "autoimmunity"[All Fields] OR "autoimmunities"[All Fields] OR "autoimmunization"[All Fields] OR "autoimmunizing"[All Fields]) AND ("muscular diseases"[MeSH Terms] OR ("muscular"[All Fields] AND "diseases"[All Fields]) OR "muscular diseases"[All Fields] OR "myopathies"[All Fields] OR "myopathy"[All Fields]) AND (("whole"[All Fields] OR "wholeness"[All Fields] OR "wholes"[All Fields]) AND ("human body"[MeSH Terms] OR ("human"[All Fields] AND "body"[All Fields]) OR "human body"[All Fields] OR "body"[All Fields]) AND ("magnetic resonance imaging"[MeSH Terms] OR ("magnetic"[All Fields] AND "resonance"[All Fields] AND "imaging"[All Fields]) OR "magnetic resonance imaging"[All Fields] OR "mri"[All Fields])) |
| Myositis AND whole body MRI | ("myositis"[MeSH Terms] OR "myositis"[All Fields] OR "myositides"[All Fields]) AND (("whole"[All Fields] OR "wholeness"[All Fields] OR "wholes"[All Fields]) AND ("human body"[MeSH Terms] OR ("human"[All Fields] AND "body"[All Fields]) OR "human body"[All Fields] OR "body"[All Fields]) AND ("magnetic resonance imaging"[MeSH Terms] OR ("magnetic"[All Fields] AND "resonance"[All Fields] AND "imaging"[All Fields]) OR "magnetic resonance imaging"[All Fields] OR "mri"[All Fields])) |
| Myositis AND dedicated MRI | ("myositis"[MeSH Terms] OR "myositis"[All Fields] OR "myositides"[All Fields]) AND (("dedicate"[All Fields] OR "dedicated"[All Fields] OR "dedicating"[All Fields] OR "dedication"[All Fields]) AND ("magnetic resonance imaging"[MeSH Terms] OR ("magnetic"[All Fields] AND "resonance"[All Fields] AND "imaging"[All Fields]) OR "magnetic resonance imaging"[All Fields] OR "mri"[All Fields])) |

| | |
|---|---|
| Inflammatory myopathies AND Magnetic resonance imaging AND localized | ("myositis"[MeSH Terms] OR "myositis"[All Fields] OR ("inflammatory"[All Fields] AND "myopathies"[All Fields]) OR "inflammatory myopathies"[All Fields]) AND ("magnetic resonance imaging"[MeSH Terms] OR ("magnetic"[All Fields] AND "resonance"[All Fields] AND "imaging"[All Fields]) OR "magnetic resonance imaging"[All Fields]) AND ("focal"[All Fields] OR "focalities"[All Fields] OR "focality"[All Fields] OR "focalization"[All Fields] OR "focalized"[All Fields] OR "focally"[All Fields] OR "focals"[All Fields] OR "local"[All Fields] OR "localisation"[All Fields] OR "localisations"[All Fields] OR "localise"[All Fields] OR "localised"[All Fields] OR "localises"[All Fields] OR "localising"[All Fields] OR "localization"[All Fields] OR "localizations"[All Fields] OR "localize"[All Fields] OR "localized"[All Fields] OR "localizer"[All Fields] OR "localizers"[All Fields] OR "localizes"[All Fields] OR "localizing"[All Fields] OR "locally"[All Fields] OR "locals"[All Fields]) |
| Dermatomyositis AND MRI | ("dermatomyositis"[MeSH Terms] OR "dermatomyositis"[All Fields]) AND ("magnetic resonance imaging"[MeSH Terms] OR ("magnetic"[All Fields] AND "resonance"[All Fields] AND "imaging"[All Fields]) OR "magnetic resonance imaging"[All Fields] OR "mri"[All Fields]) |
| MRI AND antisynthetase | ("magnetic resonance imaging"[MeSH Terms] OR ("magnetic"[All Fields] AND "resonance"[All Fields] AND "imaging"[All Fields]) OR "magnetic resonance imaging"[All Fields] OR "mri"[All Fields]) AND ("antisynthetase"[All Fields] OR "antisynthetases"[All Fields]) |
| Autoimmune myopathies AND MRI | ("autoimmune"[All Fields] OR "autoimmunity"[MeSH Terms] OR "autoimmunity"[All Fields] OR "autoimmunities"[All Fields] OR "autoimmunization"[All Fields] OR "autoimmunizing"[All Fields]) AND ("muscular diseases"[MeSH Terms] OR ("muscular"[All Fields] AND "diseases"[All Fields]) OR "muscular diseases"[All Fields] OR "myopathies"[All Fields] OR "myopathy"[All Fields]) AND ("magnetic resonance imaging"[MeSH Terms] OR ("magnetic"[All Fields] AND "resonance"[All Fields] AND "imaging"[All Fields]) OR "magnetic resonance imaging"[All Fields] OR "mri"[All Fields]) |
| Myositis AND MRI | ("myositis"[MeSH Terms] OR "myositis"[All Fields] OR "myositides"[All Fields]) AND ("magnetic resonance imaging"[MeSH Terms] OR ("magnetic"[All Fields] AND "resonance"[All Fields] AND "imaging"[All Fields]) OR "magnetic resonance imaging"[All Fields] OR "mri"[All Fields]) |

| EMBASE | |
|---|---|
| **Terms** | **Strategy** |
| Myositis AND whole body MRI | ("myositis"[MeSH Terms] OR "myositis"[All Fields] OR "myositides"[All Fields]) AND (("whole"[All Fields] OR "wholeness"[All Fields] OR "wholes"[All Fields]) AND ("human body"[MeSH Terms] OR ("human"[All Fields] AND "body"[All Fields]) OR "human body"[All Fields] OR "body"[All Fields]) AND ("magnetic resonance imaging"[MeSH Terms] OR ("magnetic"[All Fields] AND "resonance"[All Fields] AND "imaging"[All Fields]) OR "magnetic resonance imaging"[All Fields] OR "mri"[All Fields])) |
| Myositis AND dedicated MRI | ("myositis"[MeSH Terms] OR "myositis"[All Fields] OR "myositides"[All Fields]) AND (("dedicate"[All Fields] OR "dedicated"[All Fields] OR "dedicating"[All Fields] OR "dedication"[All Fields]) AND ("magnetic resonance imaging"[MeSH Terms] OR ("magnetic"[All Fields] AND "resonance"[All Fields] AND "imaging"[All Fields]) OR "magnetic resonance imaging"[All Fields] OR "mri"[All Fields])) |
| Inflammatory myopathies AND whole body MRI | ("myositis"[MeSH Terms] OR "myositis"[All Fields] OR ("inflammatory"[All Fields] AND "myopathies"[All Fields]) OR "inflammatory myopathies"[All Fields]) AND (("whole"[All Fields] OR "wholeness"[All Fields] OR "wholes"[All Fields]) AND ("human body"[MeSH Terms] OR ("human"[All Fields] AND "body"[All Fields]) OR "human body"[All Fields] OR "body"[All Fields]) AND ("magnetic resonance imaging"[MeSH Terms] OR ("magnetic"[All Fields] AND "resonance"[All Fields] AND "imaging"[All Fields]) OR "magnetic resonance imaging"[All Fields] OR "mri"[All Fields])) |
| Inflammatory myopathies AND dedicated MRI | ("myositis"[MeSH Terms] OR "myositis"[All Fields] OR ("inflammatory"[All Fields] AND "myopathies"[All Fields]) OR "inflammatory myopathies"[All Fields]) AND (("dedicate"[All Fields] OR "dedicated"[All Fields] OR "dedicating"[All Fields] OR "dedication"[All Fields]) AND ("magnetic resonance imaging"[MeSH Terms] OR ("magnetic"[All Fields] AND "resonance"[All Fields] AND "imaging"[All Fields]) OR "magnetic resonance imaging"[All Fields] OR "mri"[All Fields])) |
| Inflammatory myopathies AND Whole Body Magnetic Resonance Imaging | ("myositis"[MeSH Terms] OR "myositis"[All Fields] OR ("inflammatory"[All Fields] AND "myopathies"[All Fields]) OR "inflammatory myopathies"[All Fields]) AND (("whole"[All Fields] OR "wholeness"[All Fields] OR "wholes"[All Fields]) AND ("human body"[MeSH Terms] OR ("human"[All Fields] AND "body"[All Fields]) OR "human body"[All Fields] OR "body"[All Fields]) AND ("magnetic resonance imaging"[MeSH Terms] OR ("magnetic"[All Fields] AND "resonance"[All Fields] AND "imaging"[All Fields]) OR "magnetic resonance imaging"[All Fields])) |
| Juvenile dermatomyositis AND Whole Body MRI | ("dermatomyositis"[MeSH Terms] OR "dermatomyositis"[All Fields] OR ("juvenile"[All Fields] AND "dermatomyositis"[All Fields]) OR "juvenile dermatomyositis"[All Fields]) AND (("whole"[All Fields] OR "wholeness"[All Fields] OR "wholes"[All Fields]) AND ("human body"[MeSH Terms] OR ("human"[All Fields] AND "body"[All Fields]) OR "human body"[All Fields] OR "body"[All Fields]) AND ("magnetic resonance imaging"[MeSH Terms] OR ("magnetic"[All Fields] AND "resonance"[All Fields] AND "imaging"[All Fields]) OR "magnetic resonance imaging"[All Fields] OR "mri"[All Fields])) |

| | |
|---|---|
| Adult dermatomyositis AND localized MRI | ("dermatomyositis"[MeSH Terms] OR "dermatomyositis"[All Fields] OR ("adult"[All Fields] AND "dermatomyositis"[All Fields]) OR "adult dermatomyositis"[All Fields]) AND (("focal"[All Fields] OR "focalities"[All Fields] OR "focality"[All Fields] OR "focalization"[All Fields] OR "focalized"[All Fields] OR "focally"[All Fields] OR "focals"[All Fields] OR "local"[All Fields] OR "localisation"[All Fields] OR "localisations"[All Fields] OR "localise"[All Fields] OR "localised"[All Fields] OR "localises"[All Fields] OR "localising"[All Fields] OR "localization"[All Fields] OR "localizations"[All Fields] OR "localize"[All Fields] OR "localized"[All Fields] OR "localizer"[All Fields] OR "localizers"[All Fields] OR "localizes"[All Fields] OR "localizing"[All Fields] OR "locally"[All Fields] OR "locals"[All Fields]) AND ("magnetic resonance imaging"[MeSH Terms] OR ("magnetic"[All Fields] AND "resonance"[All Fields] AND "imaging"[All Fields]) OR "magnetic resonance imaging"[All Fields] OR "mri"[All Fields])) |
| Adult dermatomyositis AND Whole-body magnetic resonance | ("dermatomyositis"[MeSH Terms] OR "dermatomyositis"[All Fields] OR ("adult"[All Fields] AND "dermatomyositis"[All Fields]) OR "adult dermatomyositis"[All Fields]) AND ("Whole-body"[All Fields] AND ("magnetic resonance spectroscopy"[MeSH Terms] OR ("magnetic"[All Fields] AND "resonance"[All Fields] AND "spectroscopy"[All Fields]) OR "magnetic resonance spectroscopy"[All Fields] OR ("magnetic"[All Fields] AND "resonance"[All Fields]) OR "magnetic resonance"[All Fields])) |
| Autoimmune myopathies AND whole body MRI | ("autoimmune"[All Fields] OR "autoimmunity"[MeSH Terms] OR "autoimmunity"[All Fields] OR "autoimmunities"[All Fields] OR "autoimmunization"[All Fields] OR "autoimmunizing"[All Fields]) AND ("muscular diseases"[MeSH Terms] OR ("muscular"[All Fields] AND "diseases"[All Fields]) OR "muscular diseases"[All Fields] OR "myopathies"[All Fields] OR "myopathy"[All Fields]) AND (("whole"[All Fields] OR "wholeness"[All Fields] OR "wholes"[All Fields]) AND ("human body"[MeSH Terms] OR ("human"[All Fields] AND "body"[All Fields]) OR "human body"[All Fields] OR "body"[All Fields]) AND ("magnetic resonance imaging"[MeSH Terms] OR ("magnetic"[All Fields] AND "resonance"[All Fields] AND "imaging"[All Fields]) OR "magnetic resonance imaging"[All Fields] OR "mri"[All Fields])) |
| Inflammatory myopathies AND Magnetic resonance imaging AND localized | ("myositis"[MeSH Terms] OR "myositis"[All Fields] OR ("inflammatory"[All Fields] AND "myopathies"[All Fields]) OR "inflammatory myopathies"[All Fields]) AND ("magnetic resonance imaging"[MeSH Terms] OR ("magnetic"[All Fields] AND "resonance"[All Fields] AND "imaging"[All Fields]) OR "magnetic resonance imaging"[All Fields]) AND ("focal"[All Fields] OR "focalities"[All Fields] OR "focality"[All Fields] OR "focalization"[All Fields] OR "focalized"[All Fields] OR "focally"[All Fields] OR "focals"[All Fields] OR "local"[All Fields] OR "localisation"[All Fields] OR "localisations"[All Fields] OR "localise"[All Fields] OR "localised"[All Fields] OR "localises"[All Fields] OR "localising"[All Fields] OR "localization"[All Fields] OR "localizations"[All Fields] OR "localize"[All Fields] OR "localized"[All Fields] OR "localizer"[All Fields] OR "localizers"[All Fields] OR "localizes"[All Fields] OR "localizing"[All Fields] OR "locally"[All Fields] OR "locals"[All Fields]) |
| Dermatomyositis AND MRI | ("dermatomyositis"[MeSH Terms] OR "dermatomyositis"[All Fields]) AND ("magnetic resonance imaging"[MeSH Terms] OR ("magnetic"[All Fields] AND "resonance"[All Fields] AND "imaging"[All Fields]) OR "magnetic resonance imaging"[All Fields] OR "mri"[All Fields]) |
| MRI AND antisynthetase | ("magnetic resonance imaging"[MeSH Terms] OR ("magnetic"[All Fields] AND "resonance"[All Fields] AND "imaging"[All Fields]) OR "magnetic resonance imaging"[All Fields] OR "mri"[All Fields]) AND ("antisynthetase"[All Fields] OR "antisynthetases"[All Fields]) |
| Autoimmune myopathies AND MRI | ("autoimmune"[All Fields] OR "autoimmunity"[MeSH Terms] OR "autoimmunity"[All Fields] OR "autoimmunities"[All Fields] OR "autoimmunization"[All Fields] OR "autoimmunizing"[All Fields]) AND ("muscular diseases"[MeSH Terms] OR ("muscular"[All Fields] AND "diseases"[All Fields]) OR "muscular diseases"[All Fields] OR "myopathies"[All Fields] OR "myopathy"[All Fields]) AND ("magnetic resonance imaging"[MeSH Terms] OR ("magnetic"[All Fields] AND "resonance"[All Fields] AND "imaging"[All Fields]) OR "magnetic resonance imaging"[All Fields] OR "mri"[All Fields]) |
| Myositis AND MRI (2000-2022) | ("myositis"[MeSH Terms] OR "myositis"[All Fields] OR "myositides"[All Fields]) AND ("magnetic resonance imaging"[MeSH Terms] OR ("magnetic"[All Fields] AND "resonance"[All Fields] AND "imaging"[All Fields]) OR "magnetic resonance imaging"[All Fields] OR "mri"[All Fields]) |

| **COCHRANE** |
|---|
| No studies |

**Supplementary Table 2**. Search terms from Taskforce - Group 2.

| PubMed and EMBASE Search terms |
|---|
| inflammatory myopath*.mp. [mp=title, abstract, original title, name of substance word, subject heading word, floating sub-heading word, keyword heading word, organism supplementary concept word, protocol supplementary concept word, rare disease supplementary concept word, unique identifier, synonyms]<br>OR<br>dermatomyosit*.mp. [mp=title, abstract, original title, name of substance word, subject heading word, floating sub-heading word, keyword heading word, organism supplementary concept word, protocol supplementary concept]<br>OR<br>dermato-myosit*.mp. [mp=title, abstract, original title, name of substance word, subject heading word, floating sub-heading word, keyword heading word, organism supplementary concept word, protocol supplementary concept word, rare disease supplementary concept word, unique identifier, synonyms]<br>OR<br>polymyosit*.mp. [mp=title, abstract, original title, name of substance word, subject heading word, floating sub-heading word, keyword heading word, organism supplementary concept word, protocol supplementary concept]<br>OR<br>poly-myosit*.mp. [mp=title, abstract, original title, name of substance word, subject heading word, floating sub-heading word, keyword heading word, organism supplementary concept word, protocol supplementary concept]<br>OR<br>Myosit*.mp. [mp=title, abstract, original title, name of substance word, subject heading word, floating sub-heading word, keyword heading word, organism supplementary concept word, protocol supplementary concept word, rare disease supplementary concept word, unique identifier, synonyms]<br>OR<br>anti-synthetase syndrome*.mp. [mp=title, abstract, original title, name of substance word, subject heading word, floating sub-heading word, keyword heading word, organism supplementary concept word, protocol supplementary concept word, rare disease supplementary concept word, unique identifier, synonyms]<br>OR<br>antisynthetase syndrome*.mp. [mp=title, abstract, original title, name of substance word, subject heading word, floating sub-heading word, keyword heading word, organism supplementary concept word, protocol supplementary concept word, rare disease supplementary concept word, unique identifier, synonyms]<br>OR<br>necrotising myopath*.mp. [mp=title, abstract, original title, name of substance word, subject heading word, floating sub-heading word, keyword heading word, organism supplementary concept word, protocol supplementary concept word, rare disease supplementary concept word, unique identifier, synonyms]<br>OR<br>necrotizing myopath*.mp. [mp=title, abstract, original title, name of substance word, subject heading word, floating sub-heading word, keyword heading word, organism supplementary concept word, protocol supplementary concept word, rare disease supplementary concept word, unique identifier, synonyms]<br>OR<br>necrotising autoimmune myopath*.mp. [mp=title, abstract, original title, name of substance word, subject heading word, floating sub-heading word, keyword heading word, organism supplementary concept word, protocol supplementary concept word, rare disease supplementary concept word, unique identifier, synonyms]<br>OR<br>necrotizing autoimmune myopath*.mp. [mp=title, abstract, original title, name of substance word, subject heading word, floating sub-heading word, keyword heading word, organism supplementary concept word, protocol supplementary concept word, rare disease supplementary concept word, unique identifier, synonyms]<br>OR<br>necrotising auto-immune myopath*.mp. [mp=title, abstract, original title, name of substance word, subject heading word, floating sub-heading word, keyword heading word, organism supplementary concept word, protocol supplementary concept word, rare disease supplementary concept word, unique identifier, synonyms] |

| |
|---|
| necrotizing auto-immune myopath*.mp. [mp=title, abstract, original title, name of substance word, subject heading word, floating sub-heading word, keyword heading word, organism supplementary concept word, protocol supplementary concept word, rare disease supplementary concept word, unique identifier, synonyms]<br>OR<br>necrotizing auto-immune myopath*.mp. [mp=title, abstract, original title, name of substance word, subject heading word, floating sub-heading word, keyword heading word, organism supplementary concept word, protocol supplementary concept word, rare disease supplementary concept word, unique identifier, synonyms]<br>OR<br>dermatopolymyosit*.mp. [mp=title, abstract, original title, name of substance word, subject heading word, floating sub-heading word, keyword heading word, organism supplementary concept word, protocol supplementary concept word, rare disease supplementary concept word, unique identifier, synonyms] |
| **AND** |
| (MRI or WBMRI).mp. [mp=title, abstract, original title, name of substance word, subject heading word, floating sub-heading word, keyword heading word, organism supplementary concept word, protocol supplementary concept word, rare disease supplementary concept word, unique identifier, synonyms]<br>OR<br>magnetic resonance imag*.mp. [mp=title, abstract, original title, name of substance word, subject heading word, floating sub-heading word, keyword heading word, organism supplementary concept word, protocol supplementary concept word, rare disease supplementary concept word, unique identifier, synonyms] |
